# Reliable quantification of renal function from frozen blood samples

**DOI:** 10.64898/2026.06.12.26355531

**Authors:** Scott R. French, Gavin C. Culwell, Haley E. Wiskoski, Juan C. Arias, Summan Zahra, Cris E. Escareno, Emma N. Heitkamp, Anozira R. Garcia, Paola Vidana, Josue B. Quijada, Raffael Kasparov, Francesca Vitali, Edward J. Bedrick, Raza Mushtaq, Gene E. Alexander, Craig C. Weinkauf

## Abstract

**BACKGROUND:** Differences in renal function may affect Alzheimer’s disease (AD) blood biomarker levels independent of AD pathology. Although renal function was unaccounted for in foundational AD blood biomarker studies, there is potential to address this through quantification of estimated glomerular filtration rate (eGFR) from frozen serum and plasma samples. However, the validity of eGFR evaluation from long-term frozen blood samples is unknown.

**METHODS:** Adults aged 50-85 with ≥2 vascular risk factors were recruited from vascular surgery or cardiology clinics in Tucson, Arizona from 2022-2025. Individuals with creatinine assessments in point-of-care whole blood (POC-WB) and frozen serum and plasma samples using the iSTAT (Abbott) were included. eGFR was calculated using the 2021 CKD-EPI creatinine equation without race. Agreement between POC-WB and frozen blood samples was assessed using Cohen’s kappa with linear weights.

**RESULTS:** 134 participants (mean age: 72.6 ± 7.5 years, 39.6% female, 23.1% chronic kidney disease) had POC-WB eGFR available. Frozen serum and plasma samples had strong agreement with POC-WB for eGFR (Kw= 0.90-0.95, P<0.001). Pre-analytical factors had minimal effect on eGFR differences between POC-WB and frozen blood samples.

**CONCLUSIONS:** Renal function can be assessed from frozen blood samples with high consistency to POC-WB, which may be particularly relevant for interpretation of AD blood biomarkers in general aging and vascular populations who often have impaired renal function.

## 1 BACKGROUND

Alzheimer’s disease (AD) is a prevalent neurodegenerative disease that affects an estimated 8 million adults over the age of 65 in the United States.(1) Blood biomarkers have emerged as cost-effective tools to detect AD pathology prior to the onset of cognitive dysfunction.(2–7) Plasma pTau217 has emerged as the most accurate blood biomarker of AD, outperforming all other blood biomarkers in predicting AD pathology and cognitive decline and demonstrating excellent concordance with positron emission tomography (PET) and cerebrospinal fluid (CSF) AD biomarkers.(2–7)

Despite high diagnostic performance in predicting the presence of AD pathology, interpretation of AD blood biomarkers, which are primarily cleared by the kidneys, can be confounded by differences in renal function, which is commonly measured using estimated glomerular filtration rate (eGFR) derived from blood or urinary creatinine.(8–15) The Mayo Clinic Study of Aging demonstrated that the degree of elevation of plasma pTau217 in participants with chronic kidney disease (CKD), defined as eGFR <60 mL/min/1.73m^2^, is similar to the elevation seen in participants with significant beta-amyloid (Aβ) deposition.(13) Furthermore, it has been shown that 30% of CKD participants have false-positive pTau217 results compared with 7% of participants without CKD.(11) In contrast to false-positives, high renal function (low creatinine) has an independent association with false-negative pTau217 results.(14) Aside from pTau217, differences in renal function significantly affect levels of other AD biomarkers including neurofilament light (NFL), glial fibrillary acidic protein (GFAP), Aβ42, and Aβ40.(8, 9) Overall, these data highlight the need for further investigation of the influence of renal function on AD blood biomarkers.

Although growing evidence suggests that renal function affects AD blood biomarker levels, many large studies in the AD field do not account for differences in renal function.(4, 16–21) However, this limitation can potentially be addressed by post-hoc analysis of creatinine from frozen blood samples. A standard clinical evaluation of renal function based on creatinine clearance is performed by testing blood at the point-of-care (POC),(22) and the validity of testing creatinine from long-term frozen serum and plasma samples remains unclear. Prior studies on the topic were limited by small sample sizes, minimal frozen storage intervals, and the absence of fresh POC samples to act as a reference standard.(23–25) As such, we sought to establish the reliability of creatinine quantification from frozen plasma and serum samples. We did this by comparing creatinine and eGFR from POC-whole blood (POC-WB) to frozen serum and plasma samples collected from a community-dwelling older age cohort of 134 participants enriched with vascular disease.

## 2 METHODS

### 2.1 Study Design and Participants

The study participants were drawn from a cohort of 225 adults between the ages of 50-85 that were prospectively recruited from vascular surgery or cardiology clinics in Tucson, Arizona from 2022-2025 to participate in the prospective Carotids and Minds (CAM) study. Of those, 134 had planned POC-WB creatinine tested because they agreed to undergo gadolinium contrasted MRI. Individuals were eligible to participate if they had two or more vascular risk factors or diseases, including smoking (current smoker or ≥10 pack year history), hypertension, hyperlipidemia, type II diabetes mellitus, coronary artery disease, or peripheral arterial disease and/or a clinical diagnosis of carotid stenosis. Prior clinical diagnoses of dementia, major depression, neurological disorders, recent (<6 months) stroke or transient ischemic attack, end-stage renal disease (ESRD), and/or did not obtain baseline blood draw and POC-WB creatinine evaluation were exclusionary. A study selection flowchart is presented in eFigure 1.

**Figure 1:**
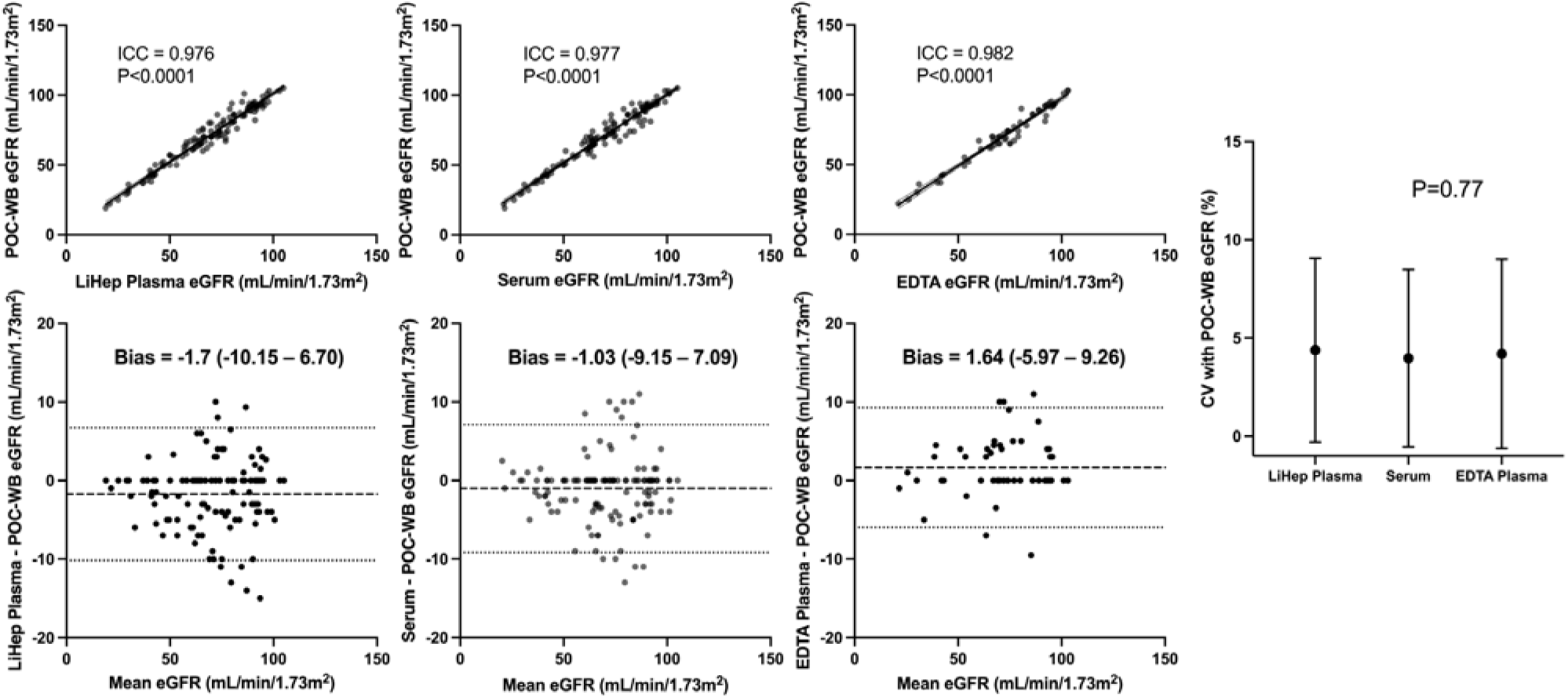
Frozen LiHep plasma, serum, and EDTA plasma eGFRs are consistent with POC-WB. Intra-class correlations and Bland-Altman plots comparing eGFR between POC-WB and frozen samples and a one-way ANOVA comparing CVs between frozen sample types is shown. Abbreviations: POC-WB, point-of-care whole blood; LiHep, lithium heparin; eGFR, estimated glomerular filtration rate; ICC, intra-class correlation; EDTA, ethylenediaminetetraacetic acid; CV, coefficient of variation.

Demographics and past medical history were determined based on participants’ self-reported responses and review of medical records. All study procedures were approved by the Institutional Review Board (1606653257) at the University of Arizona (UA), and each participant gave written informed consent after demonstrating an understanding of the study procedures.

### 2.2 Creatinine and eGFR quantification in POC-WB and frozen blood samples

Blood was drawn from 134 participants using lithium heparin (LiHep) vacutainer tubes (Becton Dickinson, Franklin Lakes, NJ, USA Cat. 367880) and clot activator tubes (Becton Dickinson, Franklin Lakes, NJ, USA Cat. 367820) and were gently inverted 10 times after filling. 59 participants also had blood drawn using ethylenediaminetetraacetic acid (EDTA) vacutainer tubes (Becton Dickinson, Franklin Lakes, NJ, USA Cat. 366643). The time between sample collection and centrifugation was recorded (pre-centrifugation interval). To obtain POC-WB eGFR, 60 μL of POC-WB was drawn directly from the LiHep vacutainer tube and tested in singlicate using the i-STAT creatinine cartridge (Abbott, Chicago, IL, USA Lots. A25147, A25191, A25349). The i-STAT creatinine cartridge is validated to assess creatinine in <3 minutes by utilizing an enzymatic reaction paired with an amperometric biosensor using POC-WB samples drawn in LiHep tubes.(22) Plasma was collected from LiHep and EDTA tubes by centrifugation at 1200 x g for 10 minutes at 25°C and serum was collected from clot activator tubes by centrifugation at 1500 x g for 15 minutes at 25°C. After centrifugation, 1 mL of supernatant was aliquoted and stored at -80°C at the UA Biorepository. POC-WB eGFR was calculated with the 2021 CKD-EPI creatinine equation without race.

To obtain creatinine values in frozen LiHep plasma, serum, and EDTA plasma samples, samples were thawed and 60 μL of sample was tested in duplicate using the i-STAT and the means were compared against POC-WB creatinine. eGFR for frozen sample types was calculated using mean creatinine values from duplicate trials with the 2021 CKD-EPI creatinine equation without race. The time spent in storage at -80°C prior to analyses and the degree of hemolysis (defined by visual assessment)(26) for each sample type was recorded. Prior to experimentation, frozen CAM samples are pulled from the UA biorepository, thawed, and re-aliquoted into 200uL and 100uL tubes to facilitate efficient in-house experimentation while minimizing wasted sample. Therefore, CAM samples typically undergo two freeze-thaw cycles prior to analysis. We set aside a subset of 52 LiHep samples that did not have multiple freeze-thaw cycles to enable exploration of freeze-thaw cycles as a variable in analyses.

### 2.3 Statistical Analysis

Intra-class correlation coefficients (ICC) and Bland-Altman plots were generated to assess agreement between eGFR obtained from POC-WB and frozen samples. Agreement between eGFR clinical categorization using eGFR from POC-WB and frozen samples was assessed using Cohen’s Kappa with linear weights (Kw). Coefficients of variation (CV) with POC-WB was compared between frozen sample types using one-way ANOVA.

To evaluate the association of hemolysis on eGFR differences between POC-WB and frozen samples, we performed Mann Whitney U tests. The effect of freeze-thaw cycles (up to three) on eGFR differences between N=52 frozen LiHep plasma and POC-WB samples was assessed using repeated-measures ANOVA. The pre-centrifugation interval and length of storage at -80°C were compared to eGFR differences between POC-WB and frozen samples using Spearman’s correlation (r_s_).

All analyses were repeated using creatinine levels. A p-value <0.05 was considered statistically significant. Data were analyzed using R Studio (2023.12.1+402) and visualized using GraphPad Prism version 10.5.0 (673) by SRF.

## 3 RESULTS

After exclusions for missing blood draw and/or POC-WB creatinine measurements (Supplemental Figure 1), the cohort consisted of 134 participants with a mean age of 72.6 ± 7.5 years and 39.6% were female. In addition, the cohort was mostly Caucasian (97.0%), 14.9% Hispanic, and contained high rates of asymptomatic carotid stenosis (56.7%) and vascular risk factors such as hypertension (75.4%), hyperlipidemia (75.4%), diabetes (30.6%), and smoking (58.2%; Table 1). POC-WB eGFR ranged from 19 mL/min/1.73m^2^ to 105 mL/min/1.73m^2^ (creatinine range 0.5 mg/dL to 3.2 mg/dL) with 31 (23.1%) participants falling below an eGFR of 60 mL/min/1.73m^2^, consistent with a clinical diagnosis of CKD. Only 9 (6.7%) participants had clinical diagnoses of CKD prior to assessing eGFR within the CAM cohort.

**Table 1:** Summary of Cohort Demographics

| Characteristic | N=134 |
| --- | --- |
| Age | 72.6 (7.5) |
| Female | 53 (39.6) |
| Education Years | 14.7 (2.6) |
| Caucasian | 130 (97.0) |
| Hispanic | 20 (14.9) |
| Obesity<sup>*</sup> | 37 (27.6) |
| Smoking<sup>†</sup> | 78 (58.2) |
| Hypertension<sup>‡</sup> | 101 (75.4) |
| Hyperlipidemia<sup>‡</sup> | 101 (75.4) |
| Diabetes<sup>‡</sup> | 41 (30.6) |
| Carotid Stenosis<sup>§</sup> | 76 (56.7) |
| Chronic Kidney Disease<sup>¶</sup> | 31 (23.1) |
| POC-WB Creatinine | 1.07 (0.39) |
| Range | 0.5–3.2 |
| POC-WB eGFR | 71.2 (19.5) |
| Range | 19–105 |
| **POC-WB eGFR Category** |  |
| ≥90 | 30 (22.4) |
| 60–89 | 73 (54.5) |
| 30–59 | 27 (20.1) |
| 15–29 | 4 (3.0) |

Continuous data are shown as mean (SD), while categorical data are shown as count (%). <sup>\*</sup>Defined as BMI ≥30. <sup>†</sup>Defined as current smoker or ≥10 pack year history.
<sup>‡</sup>Defined by the presence of a clinical diagnosis or taking associated medications.
<sup>§</sup>Defined by the presence of ≥1 carotid artery with ≥50% stenosis measured via MRI or ultrasound. <sup>¶</sup>Defined by whole blood eGFR <60 mL/min/1.73m<sup>2</sup>. Abbreviations: POC-WB, point-of-care whole blood; eGFR, estimated glomerular filtration rate.

Duplicate frozen eGFR trials demonstrated high intra-class correlation (ICC range = 0.975–0.976, P<0.0001) and low CVs (mean range: 2.12%–2.53%; Supplemental Figure 2). In addition, frozen eGFR showed high intra-class correlation with POC-WB (ICC range = 0.976–0.982, P<0.0001) and low CVs with POC-WB (Figure 1). CVs with POC-WB were equivalent between frozen sample types for eGFR (P=0.77; Figure 1). LiHep plasma tends to underestimate POC-WB eGFR by 1.7 mL/min/1.73m^2^ (bias= -1.7), with 95% of participants differing by -10.15 to 6.7 mL/min/1.73m^2^ (Figure 1). Serum tends to underestimate POC-WB eGFR by 1.03 mL/min/1.73m^2^ (bias= -1.03), with 95% of participants differing by -9.15 to 7.09 mL/min/1.73m^2^ (Figure 1). EDTA plasma tends to overestimate POC-WB eGFR by 1.64 mL/min/1.73m^2^ (bias=1.64), with 95% of participants differing by -5.97 to 9.26 mL/min/1.73m^2^ and -5.6 to 10 mL/min/1.73m^2^, respectively (Figure 1). There was strong agreement between POC-WB and frozen samples for eGFR clinical categorization (Kw= 0.90-0.95, P<0.001; Figure 2). Most of the discordance between POC-WB and frozen samples occurred in participants with eGFR >60 mL/min/1.73m^2^ (Figure 2).

**Figure 2:**
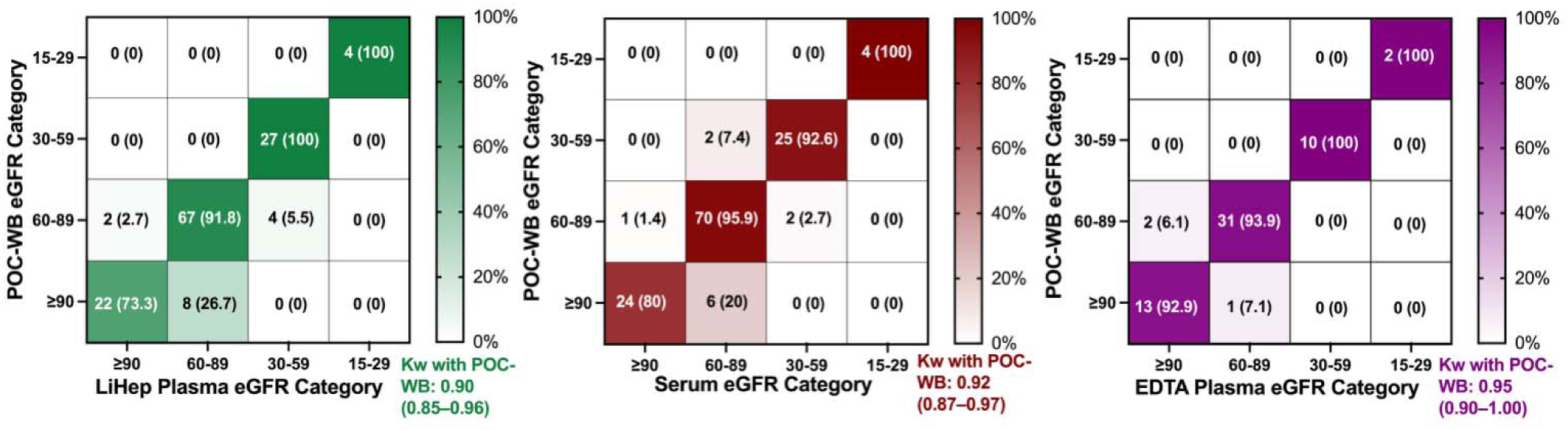
Frozen LiHep plasma, serum, and EDTA plasma have strong agreement with POC-WB for eGFR clinical categorization. Heatmaps show eGFR clinical categories assessed in frozen sample types compared to POC-WB. Agreement was assessed using Cohen’s Kappa with linear weights (kw). Abbreviations: POC, point-of-care; WB, whole blood; eGFR, estimated glomerular filtration rate; LiHep, lithium heparin; EDTA, ethylenediaminetetraacetic acid.

The pre-centrifugation interval did not correlate with eGFR differences between POC-WB and frozen LiHep plasma, serum, and EDTA plasma (LiHep plasma: rs = 0.03, P = 0.76; serum: rs = -0.04, P = 0.61; EDTA plasma: rs = -0.11, P=0.40; Figure 3). Similar results were observed for length of storage at -80°C (LiHep plasma: rs=-0.12, P=0.16; serum: rs=0.06, P=0.52; EDTA plasma: rs=-0.15, P=0.26; Figure 3). In addition, the differences in eGFR between POC-WB and LiHep plasma, serum, and EDTA plasma were unaffected by hemolysis (LiHep plasma: P=0.80; serum: P=0.70; EDTA plasma: P=0.78; Figure 4). Within a subset of 52 LiHep plasma samples, there was no significant effect of freeze-thaw cycles on eGFR differences between POC-WB and LiHep plasma (P=0.13, Supplemental Figure 3).

**Figure 3:**
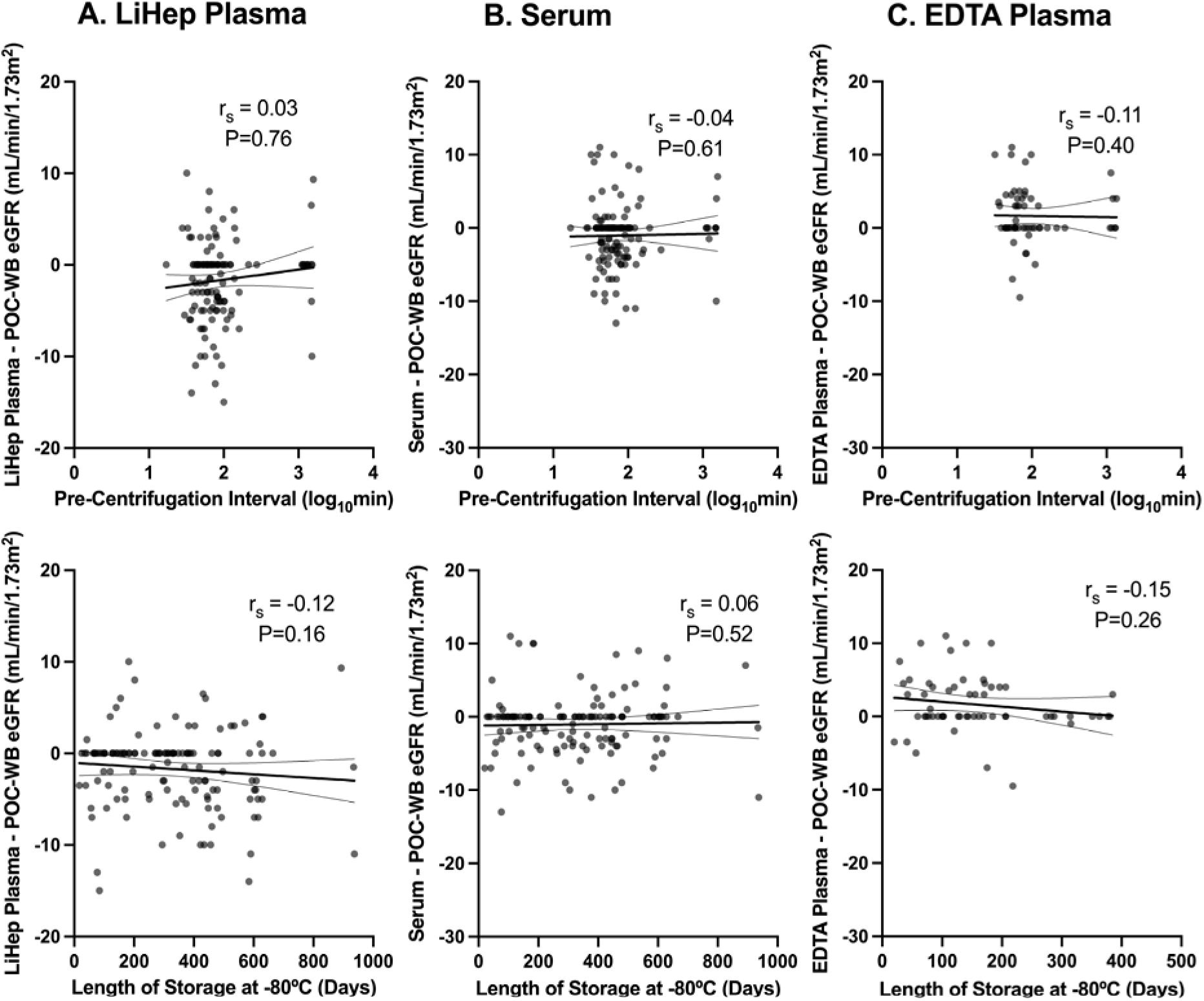
Minimal effects of storage and pre-centrifugation intervals on differences in eGFR between POC-WB and frozen samples. *Spearman’s correlations comparing differences in eGFR between POC-WB and frozen LiHep plasma, serum, EDTA plasma (N=59) to pre-centrifugation interval and length of storage. Abbreviations: POC, point-of-care; WB, whole blood; eGFR, estimated glomerular filtration rate; LiHep, lithium heparin; EDTA, ethylenediaminetetraacetic acid*.

**Figure 4:**
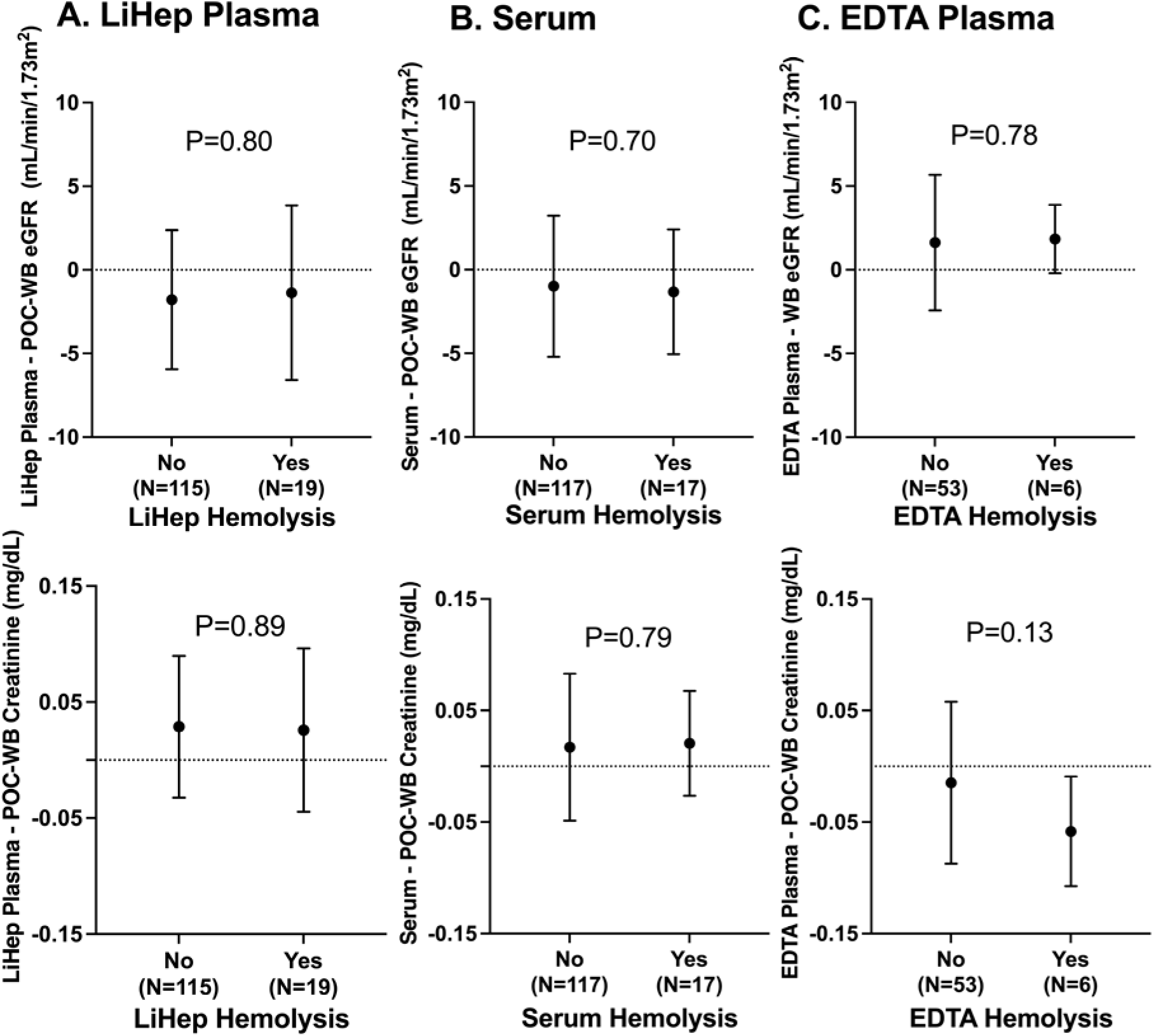
Hemolysis has minimal effects on differences in creatinine and eGFR assessed in POC-WB and frozen samples. Mann Whitney U tests show creatinine and eGFR differences between POC-WB and LiHep plasma, serum, and EDTA plasma (n=59), stratified by the presence of hemolysis. Abbreviations: POC, point-of-care; WB, whole blood; LiHep, lithium heparin; eGFR, estimated glomerular filtration rate; EDTA, ethylenediaminetetraacetic acid.

Similar to eGFR, duplicate frozen creatinine trials had high intra-class correlation and low CVs (ICC range = 0.986–0.991, P<0.0001; CV range: 2.31%–2.45%; Supplemental Figure 2). In addition, frozen creatinine had high intra-class correlation with POC-WB (ICC=0.986–0.989, P<0.0001) and low CVs (Supplemental Figure 4). CVs with POC-WB were equivalent between frozen sample types for creatinine (P=0.96; Supplemental Figure 4). LiHep plasma tends to overestimate POC-WB creatinine by 0.027 mg/dL (bias=0.027), with 95% of participants differing by -0.094 mg/dL to 0.149 mg/dL (Supplemental Figure 4). Serum tends to overestimate POC-WB creatinine by 0.017 mg/dL (bias=0.017), with 95% of participants differing by -0.107 mg/dL to 0.142 mg/dL (Supplemental Figure 4). Finally, EDTA plasma tends to underestimate POC-WB creatinine by 0.019 mg/dL (bias= -0.019), with 95% of participants differing by -0.159 mg/dL to 0.121 mg/dL (Supplemental Figure 4).

The pre-centrifugation interval did not correlate with differences between POC-WB creatinine and creatinine from LiHep plasma, serum, and EDTA plasma (LiHep plasma: rs=-0.06, P=0.53; serum: rs=0.03, P=0.77; EDTA plasma: rs=0.11, P=0.40; Supplemental Figure 5). Although a minimal effect size, longer length of storage at -80°C was associated with greater differences between LiHep plasma and POC-WB creatinine (rs=0.23, P=0.008), but this was not observed for serum or EDTA plasma (serum: rs=-0.07, P=0.42; EDTA plasma: rs=0.11, P=0.42; Supplemental Figure 5). Additionally, the differences in creatinine between POC-WB and LiHep plasma, serum, and EDTA plasma samples were unaffected by hemolysis (LiHep plasma: P=0.89; serum: P=0.79; EDTA plasma: P=0.13; Figure 4). There was no significant effect of freeze-thaw cycles on creatinine differences between POC-WB and LiHep plasma (n=52, P=0.28, Supplemental Figure 3).

## 4 DISCUSSION

In this prospectively enrolled cohort of older-aged, community-dwelling patients with vascular disease designed to study long-term dementia risk, we validated the use of frozen plasma and serum samples against fresh POC-WB (i.e., never frozen) because it is not well-established whether frozen blood samples correlate well with POC-creatinine testing. Our data demonstrate strong concordance between POC-WB and frozen serum/plasma for quantification of renal function, with minimal effects from freeze-thaw cycles, hemolysis, time to initial centrifugation, and length of storage at -80°C, which is relevant for the many aging and dementia research cohorts that have not performed POC creatinine testing.

This vascular disease cohort (mean age 72.6 years, 39.6% female) consisted of community-dwelling older adults with no prior clinical diagnoses of dementia or ESRD. As expected, this cohort had a higher prevalence of vascular comorbidities compared to the general population. Although participants with ESRD were ineligible for enrollment, CAM displayed a broad range of renal function, with 23.1% of participants meeting clinical criteria for CKD, consistent with the general older adult population.(27) This is somewhat unique compared to many other studies in the AD field, which report a much lower prevalence of CKD.(8–10, 12–14, 28). Notably, prior to measuring eGFR, only 6.7% of the CAM cohort had a previous clinical diagnosis of CKD, meaning that 70% of CKD cases would have been missed without measuring eGFR. These may help to explain the low prevalence of CKD reported in AD studies that rely on clinical diagnosis alone.(13)

Many large aging and dementia cohorts have AD blood biomarker testing available from frozen samples, but do not have available data on POC-WB eGFR/creatinine.(4, 16–21, 29) Aside from potential missed CKD diagnoses, we considered this a major limitation for AD blood biomarker analyses, and, largely because of this consideration, we investigated the validity of testing eGFR/creatinine from frozen plasma and serum samples (0.5-31 months frozen at -80°C). We found that frozen samples offer a reasonable alternative to POC-WB for evaluation of renal function, with minimal effects from freeze-thaw cycles, hemolysis, time to initial freezing and storage time at -80°C. One consideration in these analyses is that we used the i-STAT device, which is a common, validated POC machine used clinically for testing creatinine. We did not validate other creatinine testing platforms or alternative methods for calculating renal clearance, but i-STAT has demonstrated strong agreement with other common creatinine quantification platforms.(22, 30)

This study has some limitations: **1)** Individuals with ESRD are not enrolled in the CAM study, limiting the generalizability of these results to patients with ESRD. **2)** EDTA plasma samples were only available in a subset of 59 participants. However, this subset has a broad range of renal function, representative of the full CAM cohort (20.3% CKD, eGFR range: 22–103 mL/min/1.73m^2^).

In conclusion, renal function can be reliably assessed in frozen serum and plasma, producing results with high agreement and consistency to POC testing. These data provide impetus to the AD field to assess creatinine in stored blood samples, which may be particularly important as plasma pTau217 and other AD blood biomarkers are utilized in more general aging and vascular patient populations.

## Data Availability

The deidentified data utilized in the analyses for this study will be made available upon reasonable request to the corresponding author (CW) and receipt of a signed data access agreement form.

## List of Abbreviations

AD, Alzheimer’s disease; eGFR, estimated glomerular filtration rate; POC, point-of-care; WB, whole blood; PET, positron emission tomography; CSF, cerebrospinal fluid; CKD, chronic kidney disease; Aβ, beta-amyloid; NFL, neurofilament light chain; GFAP, glial fibrillary acidic protein; CAM, Carotids and Minds; ESRD, end-stage renal disease; UA, University of Arizona; LiHep, lithium heparin; EDTA, ethylenediaminetetraacetic acid; ICC, intra-class correlation; Kw, Cohen’s kappa with linear weights; CV, coefficient of variation.

## ACKNOWLEDGEMENTS

This project was supported by the NIA (R01AG070987, P30AG072980), Arizona DHS, ABRC, and the Highberger award. We thank UA CATS for assisting with blood draws and the UA Biorepository for processing and storing CAM blood samples. We also thank Wei Zhou, MD, and Steven Goldman, MD, for allowing us to store samples in their -80°C freezers prior to experimentation.

## DISCLOSURES

**Funding:** No funding source had input into study design, data analysis, or decision to publish.

**Conflicts of Interest:** None of the authors declare that they have conflicts of interest.

**Ethics Approval:** All experiments were conducted in accordance with the Declaration of Helsinki. All procedures are approved by the Institutional Review Board at the University of Arizona (1606653257) and all participants, or their legal representatives gave informed consent.

**Consent to Participate:** All subjects or their legal representatives signed written informed consent.

**Consent for Publication:** All authors listed on this manuscript are in agreement with the findings and their interpretation and provide consent for publication of this manuscript.

**Availability of data and material:** The deidentified data utilized in the analyses for this study will be made available upon reasonable request to the corresponding author (CW) and receipt of a signed data access agreement form.

**Code Availability:** All analytic software is publicly available as described in the methods section of this manuscript.

---

**Supplemental Figure 1.**
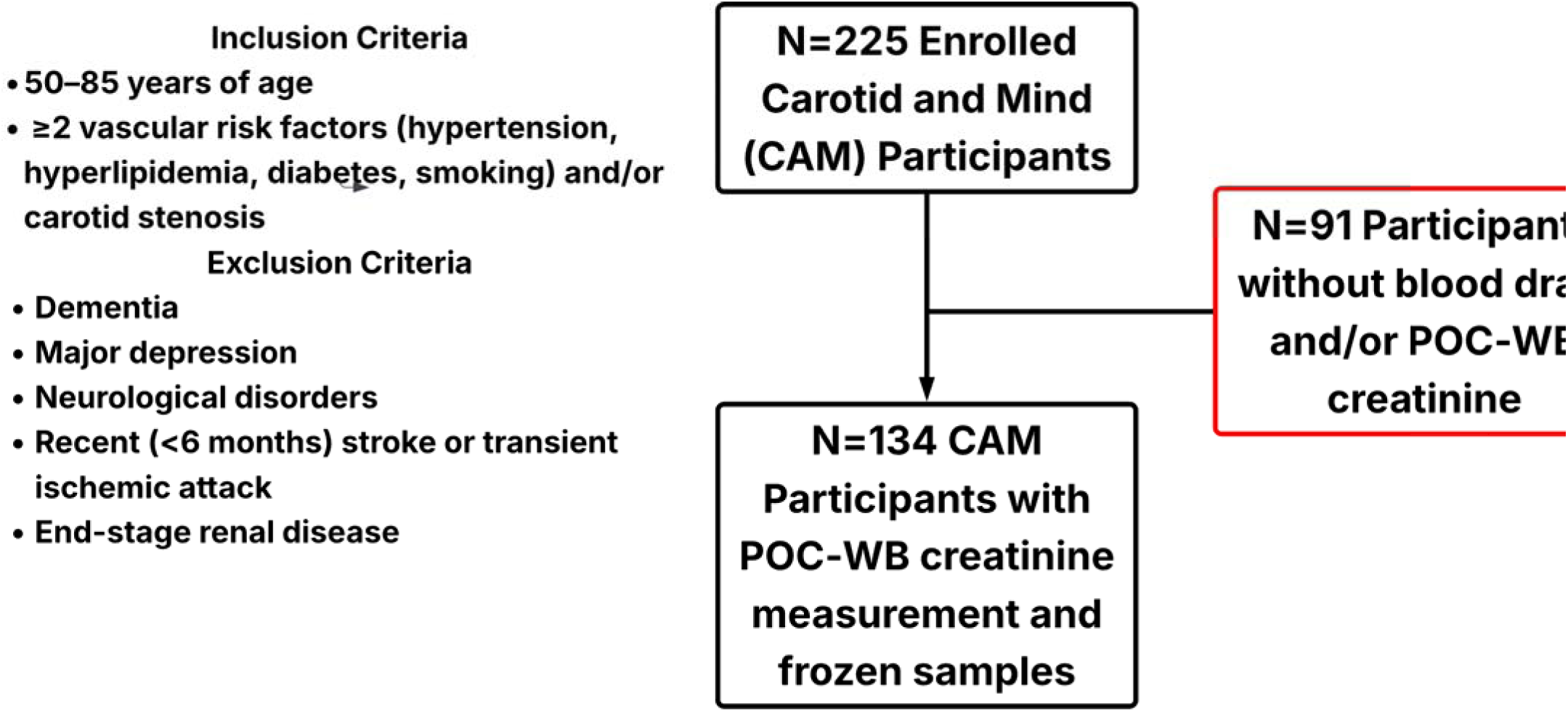
S**t**udy **selection flowchart.** The study sample selection process is shown. CAM participants were included if they had a point-of-care whole blood creatinine measurement. Abbreviations: CAM, Carotid and Mind; POC-WB, point-of-care whole blood.

**Supplemental Figure 2.**
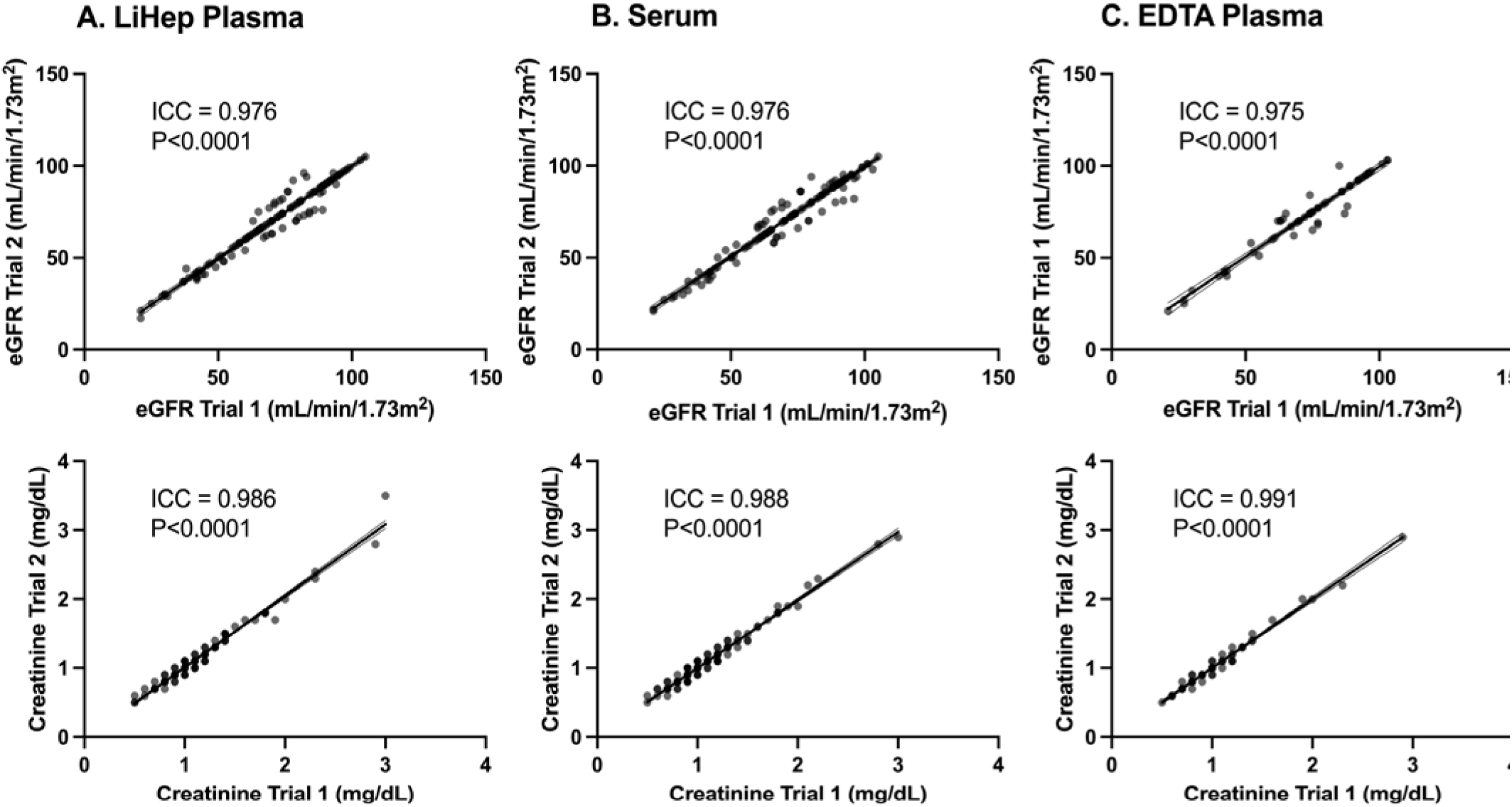
L**o**w **variation is observed between duplicate trials of creatinine and eGFR in frozen LiHep plasma, serum, and EDTA plasma.** Intra-class correlations comparing creatinine and eGFR between duplicate trials in LiHep plasma, serum, and EDTA (n=59) are shown. Abbreviations: LiHep, lithium heparin; eGFR, estimated glomerular filtration rate; EDTA, Ethylenediaminetetraacetic acid.

**Supplemental Figure 3:**
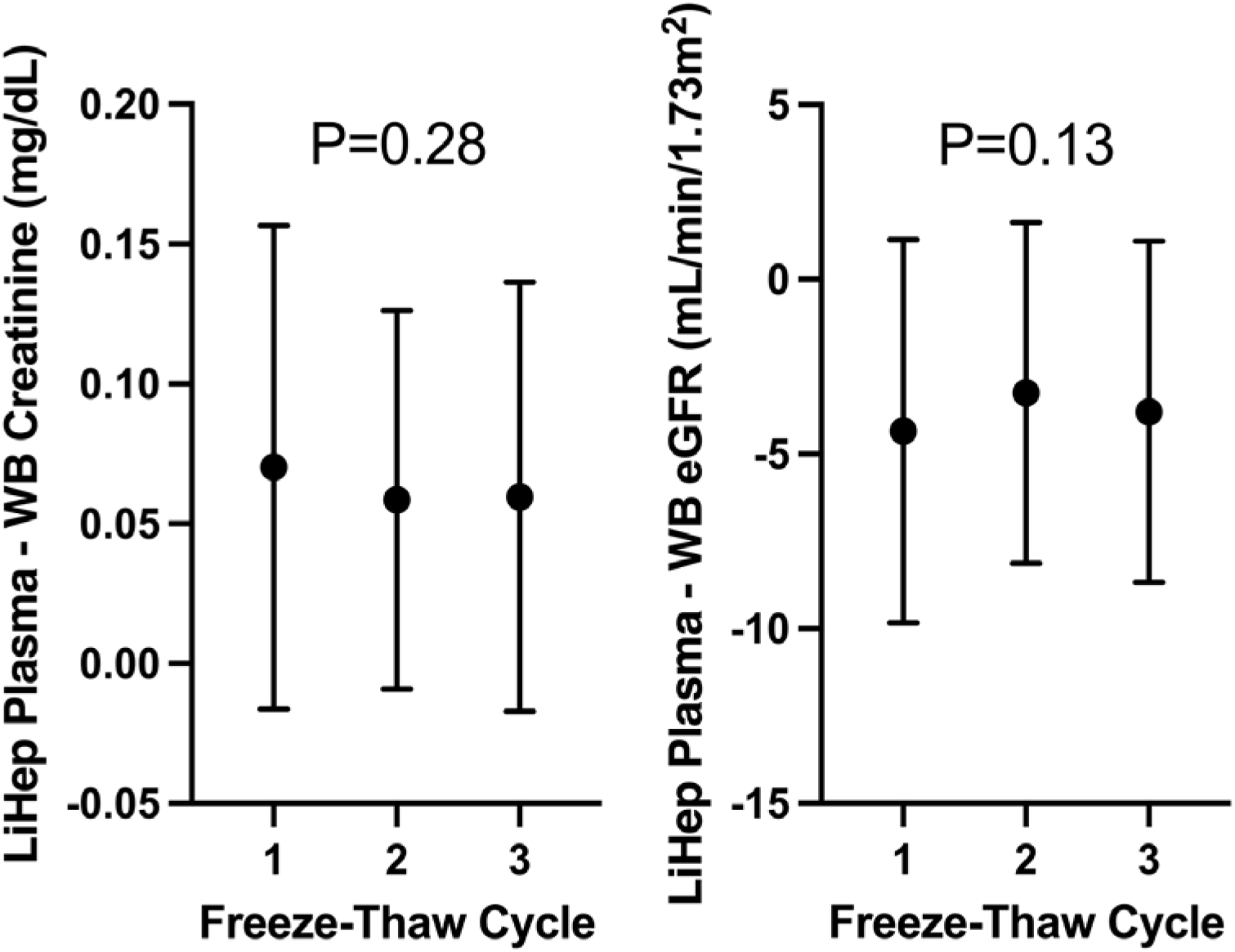
Freeze-thaw cycles have minimal effects on differences in creatinine and eGFR between frozen LiHep plasma and POC-WB in a subset of 52 participants. Repeated-measures ANOVAs comparing differences between LiHep plasma (n=52) and POC-WB creatinine and eGFR across three freeze-thaw cycles are shown. Abbreviations: POC, point-of-care; WB, whole blood; LiHep, lithium heparin; eGFR, estimated glomerular filtration rate.

**Supplemental Figure 4:**
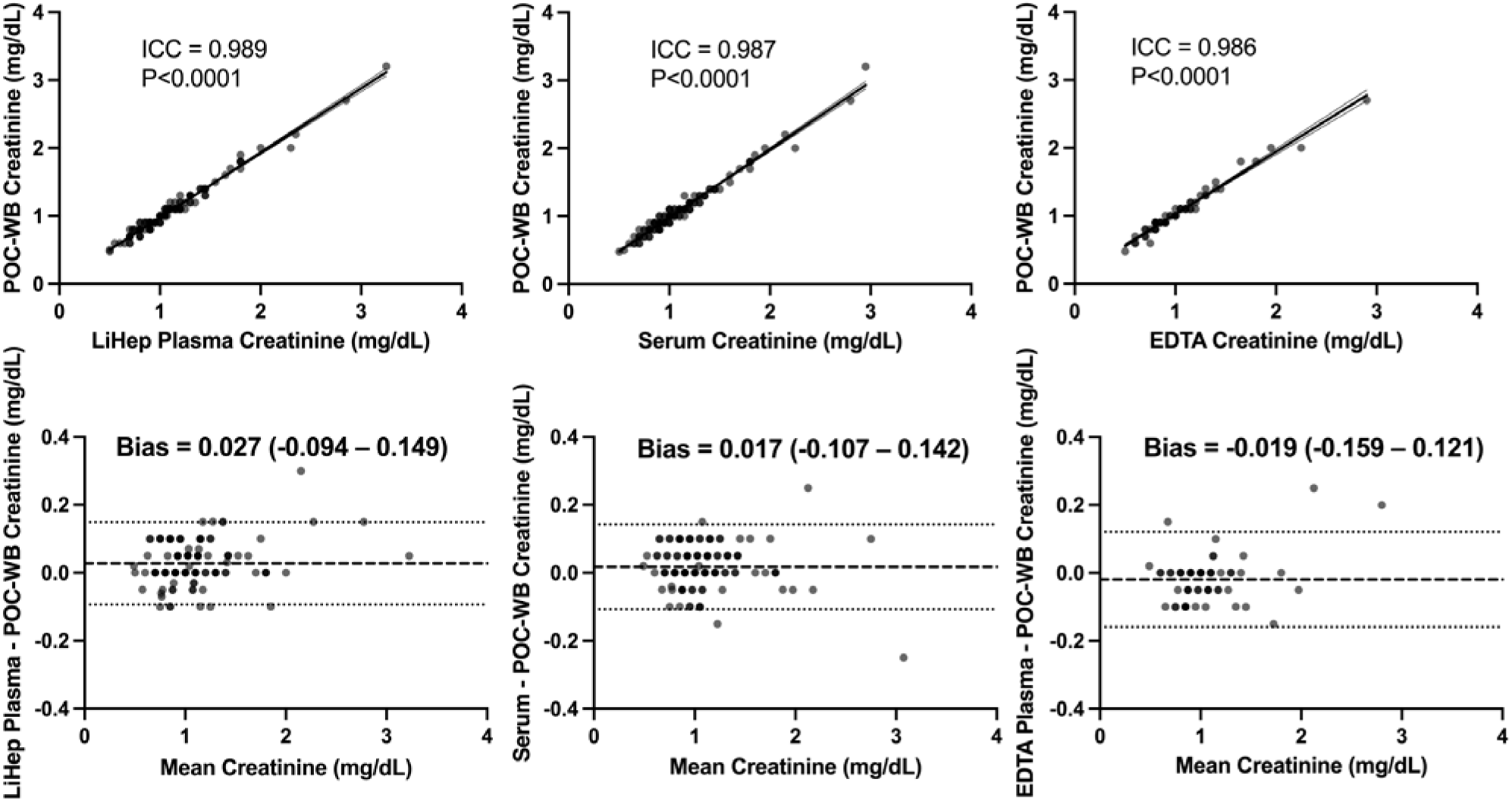
Frozen LiHep plasma, serum, and EDTA plasma creatinine levels are consistent with POC-WB. Intra-class correlations and Bland-Altman plots comparing creatinine between POC-WB and frozen samples and a one-way ANOVA comparing CVs between frozen sample types with is shown. Abbreviations: POC, point-of-care; WB, whole blood; LiHep, lithium heparin; ICC, intra-class correlation; EDTA, ethylenediaminetetraacetic acid; CV, coefficient of variation.

**Supplemental Figure 5:**
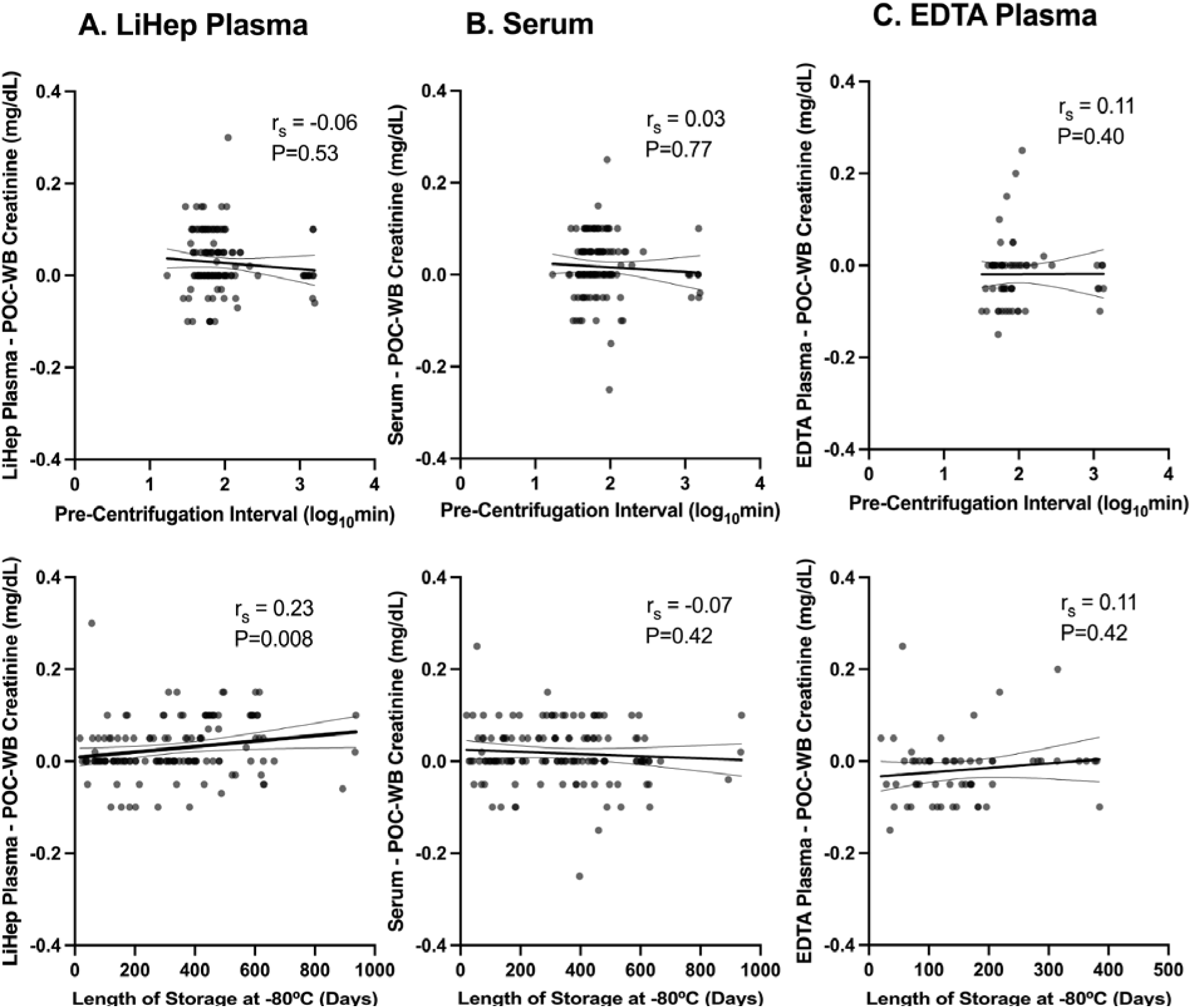
Effects of storage and sample processing intervals on differences between creatinine assessed in POC-WB and frozen samples. Spearman’s correlations comparing differences in creatinine between POC-WB and frozen LiHep plasma, serum, EDTA plasma (N=59) to pre-centrifugation interval and length of storage. Abbreviations: POC, point-of-care; WB, whole blood; LiHep, lithium heparin; EDTA, ethylenediaminetetraacetic acid.

